# Multicentre retrospective analysis of lung function impairment and risks for restrictive syndrome during childhood after type III esophageal atresia repair

**DOI:** 10.1101/2025.01.29.25321318

**Authors:** Jeanne Goulin, Thomas Brigly, Rony Sfeir, Patrick Desbordes de Cepoy, Arnaud Bonnard, Véronique Rousseau, Thomas Gelas, Audrey Guinot, Edouard Habonimana, Pascale Micheau, Aline Ranke, Isabelle Talon, Sabine Irtan, Thierry Lamireau, Pierre-Yves Rabattu, Frédéric Elbaz, Nicolas Kalfa, Nicoleta Panait, Virginie Fouquet, Hubert Lardy, Aurélien Scalabre, Philippe Buisson, Marc Margaryan, Frédéric Auber, Céline Grosos, Corinne Borderon, Cécilia Tölg, François Bastard, Françoise Troussier, Frédéric Gottrand, Françoise Schmitt

## Abstract

**Objectives:** To identify the factors that result in a restrictive ventilatory impairment during childhood following type III esophageal atresia (EA) repair.

**Study design:** A multicentre, retrospective, national cohort study was conducted on 503 patients who had undergone surgery for EA between 2008 and 2013. The results of pulmonary function tests (PFT) performed during childhood were used to compare patients with pure restrictive lung impairment to children with normal PFT. Subsequently, logistic regression was employed to ascertain potential risk factors for restrictive syndrome in type III EA.

**Results:** The cohort comprised 503 patients, of whom 216 (42.9%) had interpretable PFT. Among them, 63.4% exhibited normal results, 26.9% pure restriction, 5.1% pure obstruction, and 4.6% a mixed pattern. Patient-associated factors that were associated with a restrictive impairment were birth weight, Caucasian ethnicity (odds ratio (OR) 4.3 [1.2–15.4]), and the presence of neonatal heart defects (OR = 5.8). [1.9-16.9]), tracheomalacia (OR = 4.1 [1.6 - 10.2]) and neonatal GERD (OR = 3.1 [1.3-7.4]). The sole healthcare-associated factor was the use of respiratory crisis treatment during childhood (OR = 4.8 [1.3-18.0]), whereas neither surgical factor nor postoperative parietal thoracic deformity was associated with restriction.

**Conclusion:** In contrast to surgical approaches or chest wall abnormalities, neonatal EA-associated conditions appear to be associated with a restrictive pattern during childhood, but further prospective studies remain mandatory to validate these results.

## 1. Introduction

Esophageal atresia (EA) is a rare congenital birth defect, with an incidence of 1.9/10,000 live births [1]. This results in the formation of two separate segments of the thoracic esophagus, with the lower one being related to the trachea by a fistula in its most common anatomical form, encountered in approximately 80-85% of cases (type III EA in the Ladd Classification) [2,3]. The surgical repair of this malformation in the neonatal period involves closing the fistula and restoring esophageal continuity. Traditionally, this has been performed by a dorsolateral muscle-sparing thoracotomy. However, the thoracoscopic approach has recently emerged as a viable alternative. The latter has been demonstrated to yield comparable outcomes in terms of surgical efficacy and postoperative complications of the esophagus [4,5], with the additional benefit of causing approximately three times fewer parietal thoracic sequelae [4,6]. Previous studies of patients operated on for congenital heart diseases have indicated that a restrictive syndrome may develop in approximately 20% of cases, particularly when there have been multiple thoracic interventions and when the patient is younger [7], suggesting a potential association between parietal sequelae and respiratory impairment. The topic of pulmonary outcomes after EA has been addressed in several studies, with a prevalence of restrictive syndrome ranging from 11 to 42%. However, these studies have employed different definitions and measures, and have focused on patients of varying ages, from early childhood to adulthood [8,9]. Notably, none of these studies have specifically explored the potential role of postoperative chest wall deformities as a risk factor. There is no specific treatment for restrictive ventilatory impairment, except for the removal or prevention of its causes. Therefore, the objective of this study was to identify potential risk factors for the development of restrictive syndrome in children following surgical repair of type III EA, with a particular focus on surgical strategies and outcomes.

## 2. Patients and Methods

### 2.1 Patient Selection

A retrospective, multicentre, non-interventional cohort study was conducted in all voluntary participating pediatric surgery departments at a national level. Patients meeting the eligibility criteria were identified through the national population-based registry of all individuals born with EA in France from the CRACMO (Centre de Référence des Affections Congénitales et Malformatives de l’Oesophage, Lille, France) [1]. Subsequently, each local investigator assumed responsibility for parental information and data collection for all patients followed up at their surgical centre.

The study population comprised all patients with type III EA born between 1 January 2008 and 31 December 2013, following the provision of informed consent by their parents. Patients were excluded if they had other types of EA, “long gap” EA, died or were lost to follow-up before the age of four years, or if their parents objected to the study. Patients who did not undergo chest radiography and/or pulmonary function tests (PFT) during follow-up until the age of six to nine years were excluded from the final analysis.

This study was conducted in accordance with the principles set forth in the Declaration of Helsinki. The study was approved by the institutional ethics committee of the University Hospital Centre of Angers (n° 2019/02, 2019/01/16), declared to the French data protection authority (CNIL - Commission National de l’Informatique et des Libertés, ar190017v0) and registered in ClinicalTrial.gov (NCT04136795).

### 2.2 Data Collection and Outcome Measures

The perinatal data set comprised antenatal ultrasonographic signs of EA, gestational age, birth weight, sex, ethnic origin as recorded in medical charts because it was mandatory for PFT z-score calculation, and associated malformations linked or not to the VACTERL sequence. Prematurity was defined as the birth occurring before 37 weeks of amenorrhea (WA) and a low weight for gestational age status when the birth weight was below the 10th percentile for gestational age on the AUDIPOG curves [10]. Neonatal care included the length of intubation and hospital stay, the presence of clinical gastro-esophageal reflux (GERD) and its treatment, the necessity for gastrostomy feeding and the number and types of surgical procedures required. For each surgical intervention, the indication, the surgical approach, postoperative care (e.g., chest tube, stoma) and complications up to the age of 12 months were recorded.

The follow-up of these patients is conducted in accordance with the national protocol for diagnosis and care (PNDS, Plan National de Diagnostic et de Soins), which was revised in 2018 [11]. This protocol includes different paraclinical exams throughout childhood. Therefore, we collated data regarding the occurrence of chest radiographs, bronchoscopy, esophageal endoscopy, upper gastrointestinal (UGI) series, 24-hour pHmetry, and PFT, along with their respective conclusions. Two independent radiologists blindly analyzed each radiograph in order to diagnose chest wall abnormalities. PFT were interpreted in accordance with the 2012 Global Lung Initiative equations [12] for Functional Vital Capacity (FVC), Maximal Expiratory Volume in one second (FEV1), and the FEV1/FVC ratio. The associated z-scores were calculated using the online software (https://gli-calculator.ersnet.org/). A restrictive syndrome was defined as a FVC z-score below −1.64, with a normal FEV1/FVC z-score of ≥ −1.64. An obstructive syndrome was defined as an FEV1/FVC z-score of < 1.64, with a normal FVC z-score of ≥ −1.64. A mixed pattern was identified by both an FVC z-score and an FEV1/FVC z-score of < 1.64. The severity of respiratory impairment was assessed using a three-level system, with mild (z-score ≥ −1.64), moderate (z-score ≥ −2.5) or severe (z-score ≤ −4.0) impairment [13]. At the time of the PFT, a respiratory history and treatments were also collected. These were divided into two categories: chronic treatments, defined as pulmonary treatments ingested or inhaled on a daily basis for a period of at least three months, and acute treatments, defined as any punctual therapeutic use of medication within the previous six months. The nutritional status of the patients was evaluated using Body Mass Index (BMI) and Waterlow Index (weight/expected weight for height), with corresponding z-scores [14]. Overweight and obesity were defined in accordance with the International Obesity Task Force curves [15]. A Waterlow index below 90% indicated undernutrition, classified as severe (<70%), moderate (70 – 80%), or minor (80 – 90%). A diagnosis of GERD was made on the basis of the presence of clinical symptoms and/or the results of UGI series, 24-hour pH monitoring or endoscopy.

### 2.3 Statistics

The statistical analysis was conducted using GraphPad Prism 8.0.2 and IBM-SPSS 29.0.0.0 for Windows software. A p-value of less than 0.05 was considered statistically significant. A descriptive analysis was conducted on all data collected from the subjects included in the study. Qualitative variables were expressed in terms of frequency and percentage, while quantitative variables were expressed in terms of median and interquartile range or mean and standard deviation. A comparative analysis was conducted between patients with a restrictive syndrome or normal PFT, with only subjects presenting documented and analyzable endpoints in the perinatal period and during follow-up in childhood included. The quantitative data were compared with the two-sided Student’s t-test or the non-parametric Mann-Whitney test. Qualitative data were compared using Fisher’s exact test. A logistic regression analysis using the Cox model was conducted to identify patient-associated and health care –associated factors associated with the occurrence of a restrictive syndrome. The data included in the model were sourced from the bivariate analysis. An association was considered to be potentially significant when the p-value was less than 0.20. The final values were selected using a step-by-step descending method.

## 3. Results

### 3.1. Brief presentation of the cohort

Between 2008 and 2013, 590 patients were born with type III EA in France. Of these, 503 were eligible for this study and were recruited from the 26 pediatric surgery departments participating in the study (Figure 1). Of the total number of patients, 256 (50.9%) had not undergone PFT at the time of data collection and 31 (6.1%) had PFT whose results could not be interpreted as z-scores. Hence, only 216 (42.9%) patients had usable PFT. Please refer to Supplemental Data 1 for a comprehensive overview of the data for the entire cohort and for both patient groups.

**Figure 1:**
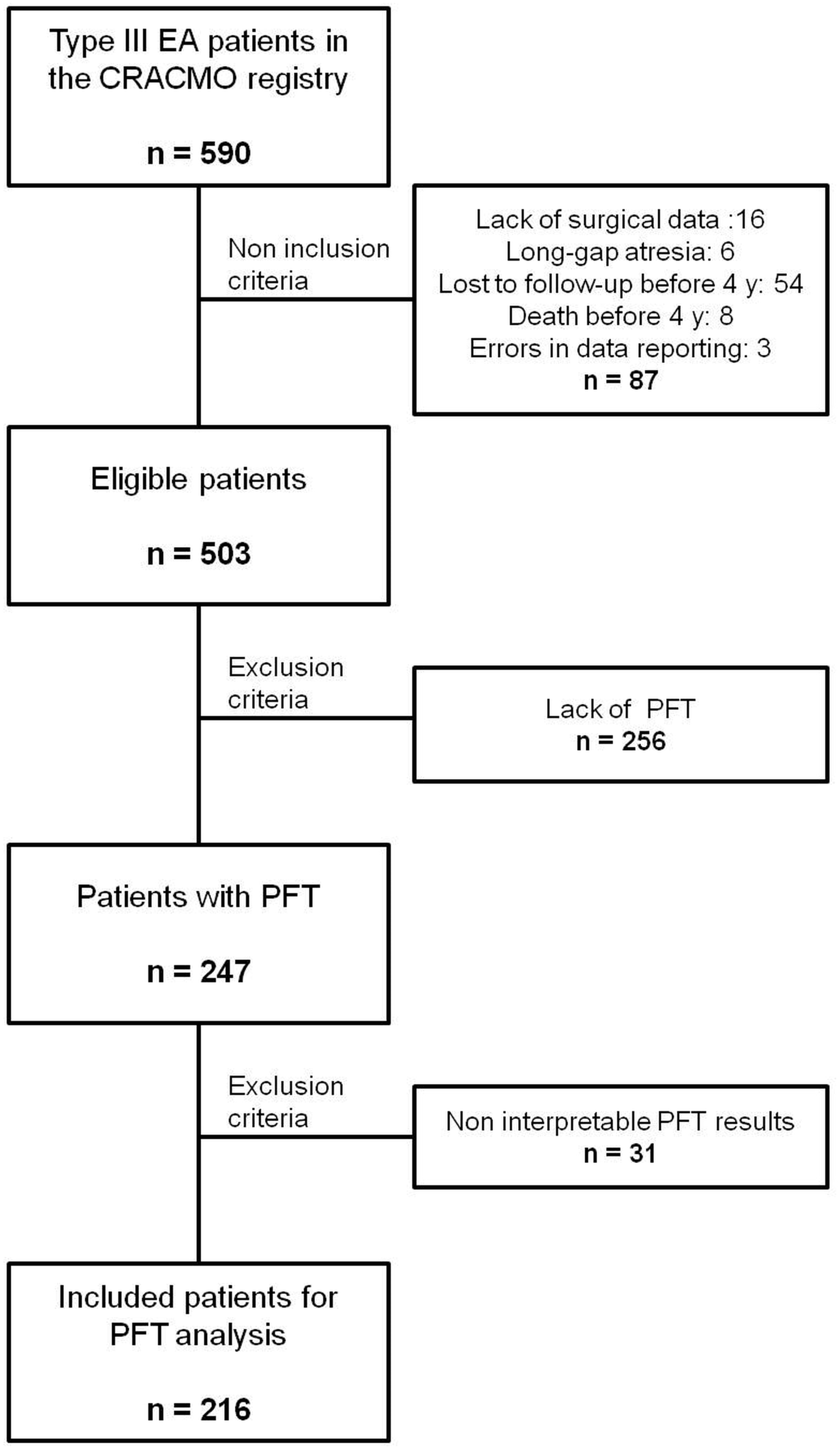
Flowchart of the screening process for patients with type III esophageal atresia (EA) included in the study of ventilatory impairment. CRACMO: Centre de Référence des Anomalies Congénitales et Malformatives de l’œsophage ; PFT: Pulmonary function tests.

### 3.2 Pulmonary function tests abnormalities

In the PFT group and at a mean age of 7.5 +/- 1.5 years, 137 (63.4%) patients exhibited normal lung function (Table 1). Impairment of ventilation was observed in 79 patients, with a restriction in 58 cases (26.9%), an obstruction in 11 cases (5.1%), and a mixed pattern in 10 cases (4.6%). The restriction patterns were classified as mild in 38 cases (55.9%), and moderate to severe in 29 cases (42.6%). Obstruction patterns were graded mild in 14 cases (66.7%) and moderate in seven cases (33.3%).

**Table 1:**
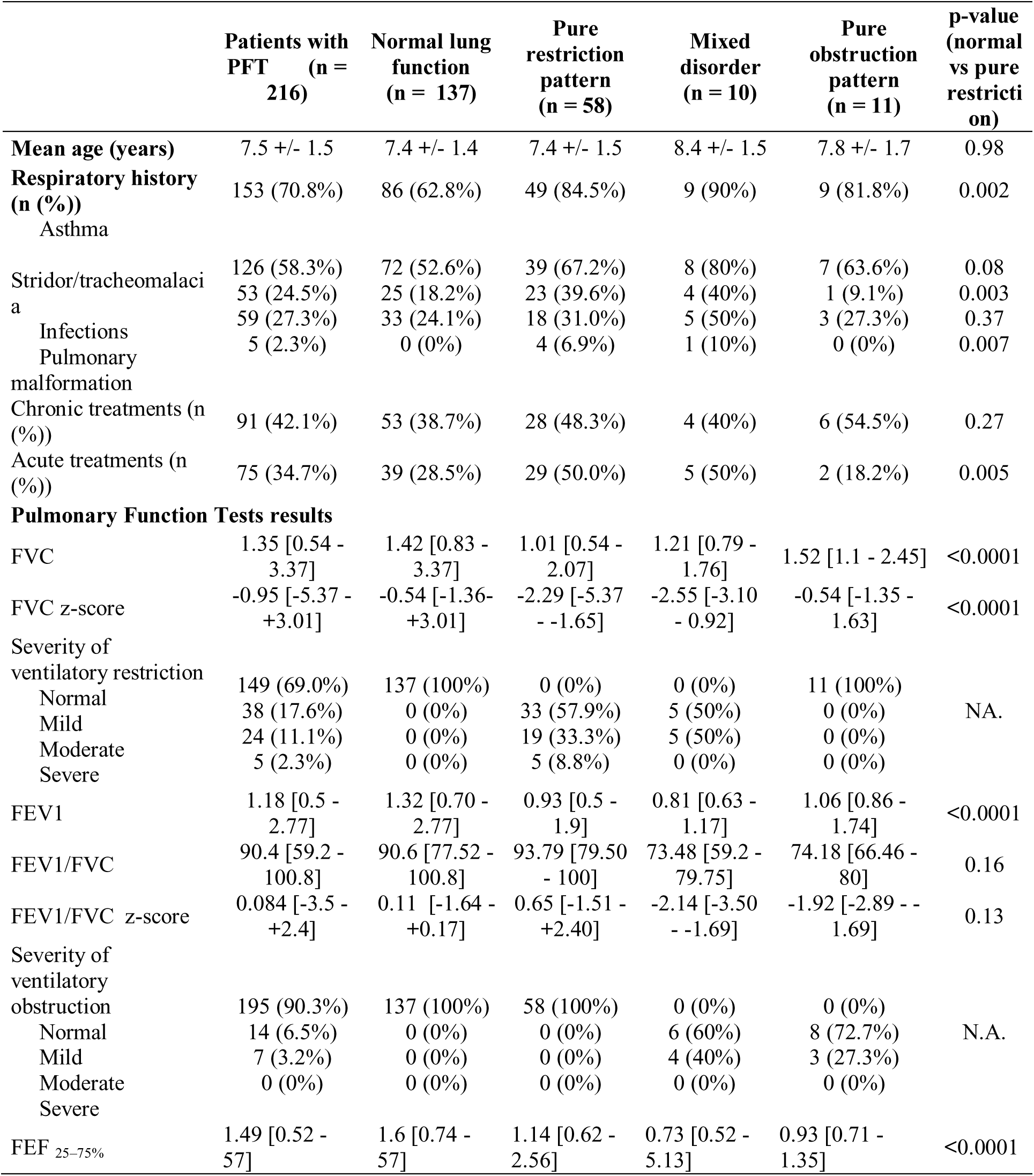
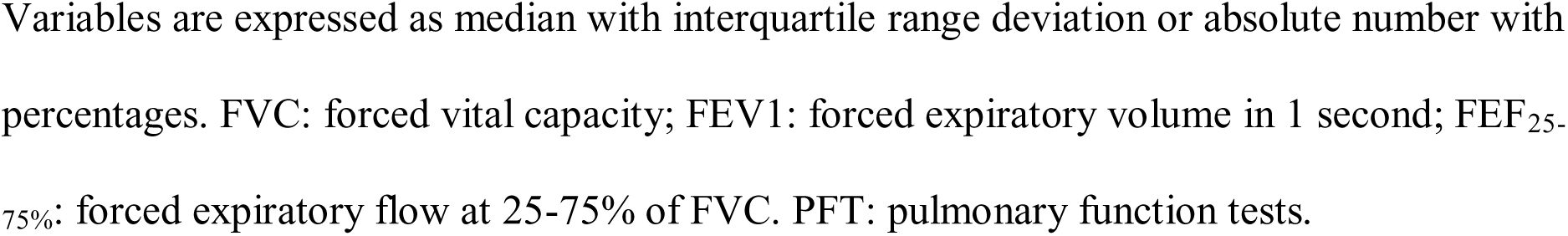
Description of respiratory history and pulmonary function tests of children with esophageal atresia repair, classified by their respiratory profiles.

Overall, 70.8% of the patients had a respiratory history, and a total of 91 patients (42.1%) required chronic treatments, comprising inhaled corticosteroids (39.4%), long-acting beta agonists (22.2%), antileukotriene agents (6.9%), and antibiotics (6.9%). Acute treatments were documented in 75 patients (34.7%), including short-acting beta agonists (23.1%), inhaled corticosteroids (10.2%), antibiotics (10.2%), and systemic corticosteroids (2.8%). Nevertheless, the impact on daily life remained low, with only two patients (0.9%) reporting partial limitations to their activities and nine (4.2%) requiring adapted schooling.

### 3.3 Comparison of patients with restriction or normal lung function

The baseline characteristics of these patients differed with regard to their birth conditions. Hence, the pure restrictive syndrome (PRS) group had 1.7 times more premature babies than the normal lung function (NLF) group. This was associated with lower gestational age and birth weight (Table 2). There was also a difference in ethnic origin, with the PRS group having 22% more Caucasian patients than the NLF group. Patients with PRS had two times more hydramnios, three times more cardiac malformations, and 2.5 times more diverse other malformations not directly related to EA.

**Table 2:** Baseline characteristics of patients with type III esophageal atresia presenting with a normal lung function or a restrictive pattern at school age.

|  | Normal lung function<br>(n = 137) | Pure restrictive<br>syndrome (n = 58) | p-value |
| --- | --- | --- | --- |
| Sex ratio | 1.8 | 1.8 | 1 |
| Ethnic origin (n (%)) |  |  |  |
| African | 23 (16.8%) | 4 (6.9%) | 0.07 |
| Caucasian | 92 (67.2%) | 50 (86.2%) | 0.008 |
| others | 9 (6.6%) | 1 (1.7%) |  |
| Birth weight (grams) | 2693 +/- 703.2 | 2330 +/- 631.2 | 0.0008 |
| Gestational age (WA) | 37.9 +/- 3.0 | 36.3 +/- 3.3 | 0.0009 |
| Prematurity | 41 (29.9%) | 30 (51.7%) | 0.005 |
| Twinning | 12/103 (11.7%) | 6/48 (12.5%) | 1 |
| Suspected antenatal diagnosis | 12 (8.8%) | 5 (8.6%) | 1 |
| Hydramnios | 39 (28.5%) | 27 (46.6%) | 0.02 |
| Associated malformations |  |  |  |
| VACTERL sequence | 25 (18.2%) | 18 (31.0%) | 0.06 |
| Cardiac malformation | 17 (12.4%) | 21 (36.2%) | 0.0003 |
| Pulmonary malformation | 2 (1.5%) | 0 | 1 |
| Other malformations | 11 (8.0%) | 12 (20.7%) | 0.016 |

Variables are expressed as mean +/- standard deviation or absolute number with percentages. WA:
weeks of amenorrhea. VACTERL sequence: malformative condition associating at least three
malformations among the vertebral column, the ano-rectal system, the heart, the tracheo-
esophageal tractus, the kidneys and the limbs.

No significant differences were observed in the surgical management between the two groups (Table 3), including the surgical approach (thoracotomy vs thoracoscopy), the timing of trachea-esophageal closure and of esophageal continuity restoration, chest tube drainage, and the occurrence of concomitant surgeries. However, gastrostomy tube insertion was more prevalent in the PRS group, occurring in 10 (17.2%) cases compared to 2 (1.5%) in the NLF group (p = 0.0001). The PRS group saw a higher proportion of inborn newborns, with longer stays in the neonatal care unit and an additional day of invasive ventilation. Furthermore, they underwent a greater number of iterative thoracic surgeries for both EA (15.5% vs 5.8%, p = 0.048) and cardiac or neuro-orthopedic concerns (17.2% vs 5.1%, p = 0.01), including more sternotomies (6 vs 0 cases, p = 0.0005), as well as more abdominal surgeries (48.3% vs 29.9%, p = 0.02). Upon discharge, 80% of patients in both groups received proton pump inhibitors for GERD treatment.

**Table 3:** Comparative analysis of neonatal and surgical cares of patients with type III esophageal atresia presenting with a normal lung function or a restrictive ventilatory impairment at school age.

|  | Normal lung<br>function<br>(n = 137) | Pure restrictive<br>syndrome (n = 58) | p-value |
| --- | --- | --- | --- |
| Inborn | 77 (56.2%) | 42 (72.4%) | 0.037 |
| Age at EA surgery (days) | 1 [0 - 11] | 1 [0 - 6] | 0.08 |
| Direct restoration of esophageal continuity | 133 (97.1%) | 52 (89.7%) | 0.067 |
| **Surgical approach** |  |  |  |
| posterolateral thoracotomy | 133 (97.1%) | 55 (94.8%) | 0.45 |
| thoracoscopy | 4 (2.8%) | 3 (5.2%) | 0.43 |
| conversion of thoracoscopy to open | 1 (0.7%) | 2 (3.4%) | 0.21 |
| Chest tube insertion | 132 (96.4%) | 55 (94.8%) | 0.7 |
| Gastrostomy insertion | 2 (1.5%) | 10 (17.2%) | 0.0001 |
| **Associated surgery (during EA repair)** | 9 (6.6%) | 6 (10.3%) | 0.39 |
| Cardiac | 1 (0.7%) | 0 | 0.06 |
| Low ARM repair | 2 (1.5%) | 4 (6.9%) | 0.06 |
| High ARM repair | 3 (2.2%) | 3 (5.2%) | 0.36 |
| Colostomy | 6 (4.4%) | 3 (5.2%) | 0.73 |
| Duodenal atresia | 3 (2.2%) | 4 (6.9%) | 0.2 |
| Other | 1 (0.7%) | 7 (12.1%) | 0.001 |
| Length of invasive ventilation (days) | 3 [0 - 37] | 4 [0 - 84] | 0.0012 |
| Age at discharge (days) | 19 [5 - 380] | 30 [11 - 160] | 0.002 |
| **Second thoracic surgery for EA** | 8 (5.8%) | 9 (15.5%) | 0.048 |
| Fistula reopening | 4 (2.9%) | 3 (5.2%) | 0.43 |
| Delayed esophageal repair | 2 (1.5%) | 5 (8.6%) | 0.025 |
| Anastomotic leakage | 1 (0.7%) | 2 (3.4%) | 0.21 |
| **Other thoracic surgery** | 7 (5.1%) | 10 (17.2%) | 0.01 |
| Age at thoracic surgery (months) | 5.9 [3.5 - 72.8] | 4 [0.8 - 28.1] | 0.12 |
| Cause of thoracic surgery |  |  |  |
| Cardiac surgery | 0 (0%) | 8 (13.8%) | <0.0001 |
| Aortopexy | 3 (2.2%) | 1 (1.7%) | 1 |
| Closure of arterial canal | 1 (0.7%) | 1 (1.7%) | 0.5 |
| Neuro-orthopedic issue | 3 (2.2%) | 0 (0%) | 0.56 |
| Surgical approach |  |  |  |
| Sternotomy | 0 (%) | 6 (10.3%) | 0.0005 |
| Right posterolateral thoracotomy | 3 (2.2%) | 2 (3.4%) | 0.63 |
| Thoracoscopy | 2 (3.4%) | 0 (0%) | 1 |
| **Abdominal surgery** | 41 (29.9%) | 28 (48.3%) | 0.02 |
| Age at abdominal surgery (months) | 3 [0.3 - 93] | 2.5 [0.3 - 276] | 0.2 |
| Iterative abdominal surgery | 15 (10.9%) | 15 (25.9%) | 0.015 |

Variables are expressed as median with interquartile range or absolute number with
percentages EA: esophageal atresia; ARM: anorectal malformation.

Complications during the first year of life (Table 4) were more frequent in the PRS group (34.5% vs 18.9%, p = 0.026), predominantly due to anastomotic strictures necessitating esophageal dilations (29.3% vs 14.6%, p = 0.027). Clinical follow-up during childhood demonstrated that patients in the PRS group exhibited a higher prevalence of respiratory history, including tracheomalacia and pulmonary malformations, and a greater need for crisis treatments compared to NLF patients (Table 1). Additionally, the mean BMI z-scores and Waterlow index were lower too, resulting in twice the incidence of undernutrition (Table 4), without association with the presence of GERD. Overall, the results of paraclinical examinations, including fibroscopy, 24-hour pHmetry, esophageal series and chest radiographs, were comparable between the two groups. Chest radiographs revealed the presence of surgical-related chest wall abnormalities in 70% of cases, predominantly involving intercostal space abnormalities (88 patients, 58.3%) and costal hypoplasia (77 patients, 51.0%).

**Table 4:** Comparative analysis of survey data for children with type III esophageal atresia presenting with a normal lung function or a restrictive pattern at school age.

|  | Normal lung function<br>(n = 137) | Pure restrictive<br>syndrome (n = 58) | p-value |
| --- | --- | --- | --- |
| **First year outcomes** |  |  |  |
| Complications and management | 26 (18.9%) | 20 (34.5%) | 0.026 |
| Symptomatic anastomotic stricture* | 20 (14.6%) | 17 (29.3%) | 0.027 |
| Esophageal dilation | 42 (30.7%) | 26 (44.8%) | 0.07 |
| Fundoplication | 9 (6.6%) | 8 (13.8%) | 0.16 |
| Gastrostomy | 5/100 (5.0%) | 10/35 (28.6%) | 0.0005 |
| BMI at 6 months | 16.3 [10.7 - 39.1] | 16.1 [12.5 - 42.4] | 0.64 |
| BMI at 12 months | 16.3 [12.3 - 20.3] | 15.8 [13.1 - 19.5] | 0.23 |
| Bronchofibroscopy | 39 (28.5%) | 18 (31.0%) | 0.73 |
| Age (days) | 45 [1.2 - 2745] | 64 [1.3 - 2892] | 0.9 |
| Results |  |  |  |
| Normal | 3 (8.1%) | 4 (22.2%) | 0.2 |
| Tracheomalacia | 28 (75.7%) | 10 (55.6%) | 0.2 |
| Tracheal compression | 7 (18.9%) | 3 (16.7%) | 1 |
| **School age children nutritional status** |  |  |  |
| BMI (Kg.m<sup>-2</sup>) | 15.1 [2.1 - 26] | 14.3 [12.2 - 25] | 0.003 |
| BMI z-score | -0.18 [-2.93 - +4.40] | -0.97 [-3.16 - +4.09] | 0.0008 |
| Normal weight | 111 (81.0%) | 43 (74.1%) | 0.34 |
| Thinness | 9 (6.6%) | 11 (19.0%) | 0.017 |
| Overweight/obese | 17 (12.4%) | 4 (6.9%) | 0.32 |
| Waterlow index (%) | 100 [75 - 139] | 93.7 [76-158] | 0.009 |
| Undernutrition | 27 (19.7%) | 23 (39.7%) | 0.007 |
| **School age children paraclinic exams** |  |  |  |
| Upper gastrointestinal series | 37 (27.0%) | 16 (27.6%) | 1 |
| Age (years) | 6 [0.005 - 11] | 7 [1 - 9.6] | 0.31 |
| Results |  |  |  |
| Normal | 19 (51.4%) | 6 (37.5%) | 0.39 |
| Esophageal diameter disparity | 11 (29.7%) | 2 (12.5%) | 0.30 |
| GERD | 8 (21.6%) | 3 (18.8%) | 1 |
| Stasis | 2 (5.4%) | 1 (6.3%) | 1 |
| Diameter disparity + stasis | 1 (2.7%) | 3 (18.8%) | 0.08 |
| 24 hour pHmetry | 87 (63.5%) | 40 (69.0%) | 0.51 |
| Age (years) | 2.9 [0.3 - 57] | 3.3 [0.5 - 9] | 0.74 |
| GERD | 37 (42.5%) | 17 (42.5%) | 1 |
| Chest radiograph | 114 83.2%) | 37 (63.8%) | 0.005 |
| Age (years) | 5.8 [0.01 - 11] | 7 [0.1 - 13] | 0.1 |
| Surgery related abnormalities | 79/114 (69.3%) | 27/37 (73.0%) | 0.84 |
| Costal fusion | 6 (5.3%) | 4 (10.8%) | 0.26 |
| Intercostal space abnormality | 65 (57.0%) | 23 (62.2%) | 0.7 |
| Costal hypoplasia | 59 (51.8%) | 18 (48.6%) | 0.85 |
| Other costal abnormalities | 0 (0%) | 1 (2.7%) | 0.06 |
| Scoliosis | 14 (12.3%) | 3 (8.1%) | 0.76 |

Variables are expressed as median with interquartile range or absolute number with
percentages BMI: body mass index; GERD: gastro-oesophageal reflux disease. \* symptomatic anastomotic strictures here concern the ones occurring after neonatal unit discharge.

### 3.4 Risk factors for a restrictive syndrome at school age

The potential risk factors for a restrictive syndrome were classified into two categories: constitutional or patient-associated factors, and acquired factors, related to the medical and surgical care provided (Table 5).

**Table 5:**
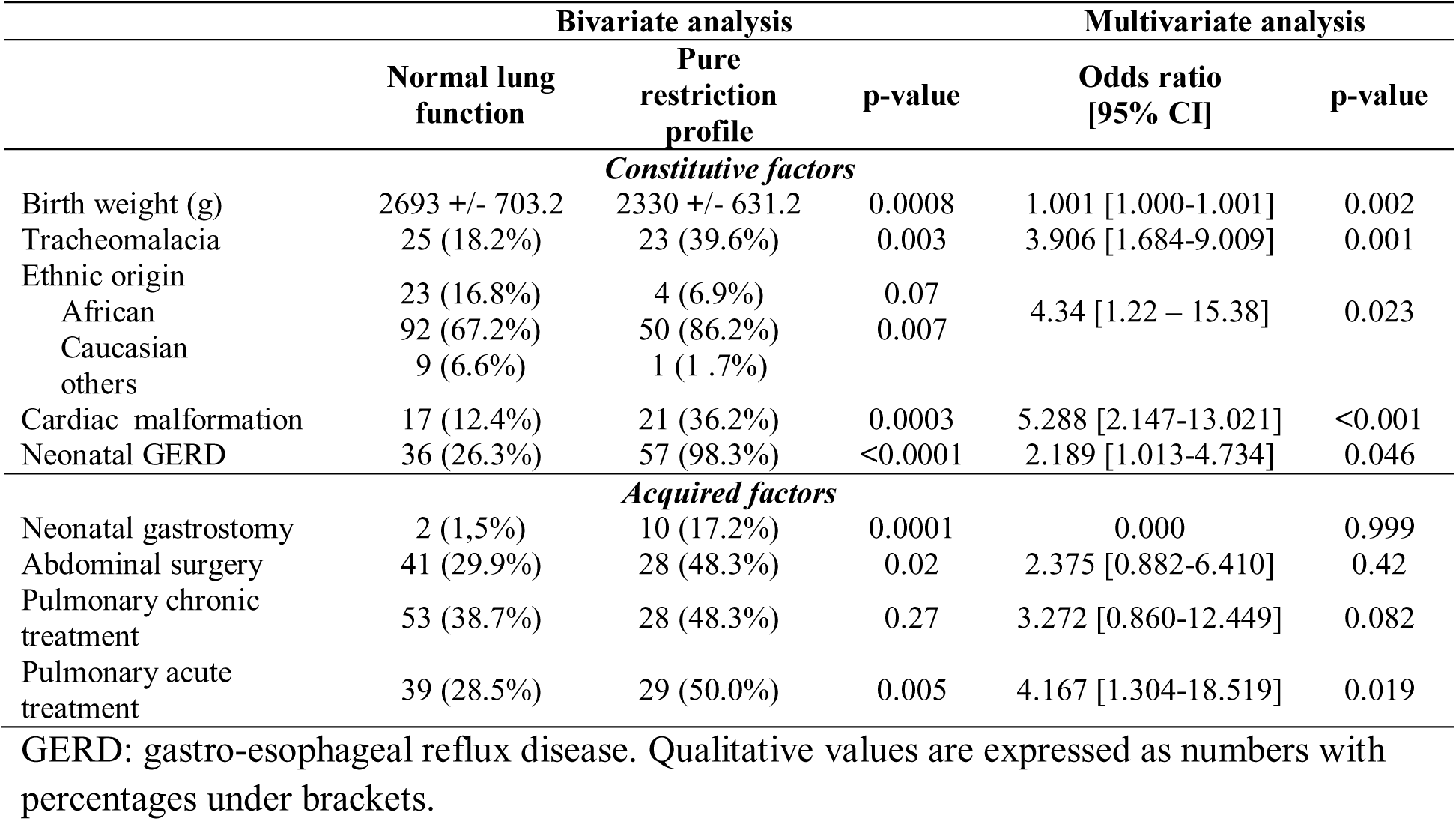
Factors associated with the occurrence of a restrictive pattern at 6 – 9 years.

The constitutional factors included in the analysis were prematurity, birth weight, ethnic origin, associated malformations from the VACTERL sequence, cardiac malformations, associated duodenal atresia, hydramnios, neonatal GERD, asthma, tracheomalacia, pulmonary infections and the Waterlow index at school age. Among the identified factors, low birth weight, Caucasian ethnicity, cardiac malformations, neonatal GERD and tracheomalacia were found to be positively associated with PRS.

The acquired factors were as follows: age at discharge home, mechanical ventilation length, initial gastrostomy insertion, iterative thoracic surgery for EA and for other causes, esophageal dilation for anastomotic stricture, and abdominal surgery during infancy, fundoplication, costal abnormalities, and pulmonary acute and chronic treatments. The sole factor identified as being associated with a restrictive syndrome was the need for acute treatment within the past six months. There was no correlation between the occurrence of respiratory impairment and thoracic and abdominal surgeries and their surgical approaches, GERD and fundoplication, as well as postoperative complications affecting the esophagus and chest wall.

## 4. Discussion

National guidelines recommend that patients with type III EA undergo multidisciplinary evaluations at school age to screen for late digestive and respiratory complications [11]. The most common respiratory symptoms include recurrent bronchopulmonary infections, wheezing, cough and asthma. However, infraclinic ventilatory impairment can only be detected by the use of PFT. Karmaus *et al.* [16] recently demonstrated that PFT values in children may be indicative of lung function and decline in adulthood. An analysis of the results according to the GLI equations [12] revealed that up to 31.5% of the children exhibited restrictive impairment, with 42.6% of cases graded as moderate to severe.

A comparative analysis of PRS and NLF patients revealed significant differences between the two populations with regard to both baseline characteristics and clinical presentation at school age. These differences often resulted in more pronounced medical and surgical needs during infancy and early childhood. It is noteworthy that no technical surgical factor appeared to have an impact on PFT abnormalities. As previously described, thoracotomy resulted in three times more chest wall deformities than thoracoscopy in an ancillary case-control study on surgical approaches in our population [17]. However, their respective frequency did not differ between the PRS and NLF groups, and thoracic parietal sequelae were not associated here with restriction. The insufficient number of thoracoscopies performed in our series precludes us from establishing a pathophysiological link between surgical approach, surgical-induced chest wall asymmetry and their potential impact on FVC by decreasing chest expansion during breathing [18]. Nevertheless, recent studies have investigated the impact of surgical approaches to lobectomy for congenital pulmonary malformations on subsequent lung function, concluding that there is no significant difference in long-term PFT results [19,20].

In this study, the first notable difference was the higher number of premature babies, along with the lower birth weight and potentially the higher number of inborn babies. These factors are known to be associated with EA, often complicating its postoperative course [21,22]. However, they have also been linked to respiratory impairment, notably by a reduction of the FVC [23–25], resulting from pulmonary hypodysplasia due to a lack of intrauterine growth and maturation. These structural abnormalities persist following EA repair, and their repercussions may be exacerbated when associated with two factors that are particularly prevalent in EA patients and identified in this study as risk factors for restrictive syndrome. The first one is tracheomalacia, leading in a decrease of bronchial clearance [26], which is present in 24 to 78% of patients [8,27,28] and has been reported here to persist during childhood in 40% of our PRS patients. The second factor is neonatal GERD, which induces chronic continuous silent aspiration and chronic lung inflammation. This may lead to recurrent pulmonary infections and fibrosis [29–31], as well as increasing tracheomalacia [32].

A new factor identified as being associated with an increased risk of restrictive syndrome in this study was ethnic origin. Patients of Caucasian ethnicity were found to have a fourfold increased risk in comparison to those of African ethnicity. In contrast to previous interpretations, the GLI-2012 equations are race- and ethnicity-adjusted [12,13]. Despite ongoing debate regarding the scientific rationale for this approach [36], recent comparative studies have demonstrated that previous equations tended to underestimate FVC and FEV1 values in African populations [12,33]. It is therefore possible that the identification of this new risk factor for restrictive syndrome is a result of the change in calculations compared to previous studies on ventilatory impairment in EA [8,28,34,35]. It will thus be important to take into account future studies on the impact of ethnicity on the interpretation of PFT before validating this result.

A further notable distinction between PRS and NLF patients was the higher prevalence of associated malformations. The most prevalent were cardiac malformations, which were present in some cases as part of the VACTERL sequence. Their association with a restrictive impairment was confirmed on multivariate analysis, with an odds ratio of 5.79. The influence of these factors may be attributed to various factors, including an impact on lung blood circulation, their association with inspiratory muscle dysfunction and elevation of inflammation markers, as previously stated by Spiesshoefer et al. [36]. Additionally, the role of surgical interventions, which have the potential to multiply the number of thoracic surgeries, must be considered. The latter has been identified as a cause of restrictive syndrome in the literature [7,37], but this was not confirmed in our series, despite the number of thoracic openings being higher in the PRS group.

In the long term, a respiratory history was identified in 70% of the patients, with half of them still requiring chronic and/or acute treatments. The relatively low prevalence of obstructive and mixed disorders may be attributed to the substantial use of these treatments. The PRS group experienced more complications associated with oesophageal anastomosis, and required initial enteral feeding via gastrostomy more frequently, but this was not directly related with further poor overall nutritional status during childhood. It is notable that undernutrition was two times more prevalent in the PRS group. However, in two-thirds of cases, it was classified as minor, indicating that nutritional support during childhood was generally satisfactory. Undernutrition and stunting have been identified in 15-20% of infants born with EA [38], as well as in 20% of older children [39]. These conditions are associated with a lower resting metabolic rate and appear to persist into adulthood [40]. Furthermore, undernutrition is also encountered in severe chronic respiratory diseases [41], which are linked to hypercatabolism and hypoxemia. This may in turn have a negative impact on the mechanical function of the lung and of the inspiratory muscles [42]. These results therefore provide strong support for the systematic detection and prevention of both malnutrition and respiratory abnormalities on a lifelong medical survey of these patients.

The retrospective design and the multiplicity of surgical centres represent some of the limitations of our study. Despite the standardisation of EA care by national recommendations, some heterogeneity in medical care and potential centre effects on surgery, outcomes and routine examinations prescription remained. A potential selection bias may have been introduced, as demonstrated by the observation that patients who underwent PFT also had a greater number of other examinations. Unfortunately, it was not possible to ascertain whether this reflected more rigorous follow-up strategies or the fact that these patients had more severe outcomes. Moreover, the utilisation of PFT necessitated the inclusion of patients born in the early 2010s, where the use of thoracoscopy in this context remained relatively low. Thus, the number of patients was insufficient to draw conclusions regarding the impact of minimally invasive surgery on pulmonary function in contrast to the demonstrated outcomes in cases of chest wall deformities. The principal strengths of our study can be identified as follows: firstly, the national CRACMO registry guarantees the exhaustive screening of patients and the capacity to work with a large cohort, thereby enhancing the statistical power of the analysis; secondly, the standardisation of multidisciplinary care provided in specialised centres across the territory. The latter, in conjunction with the selection of type III EA patients, excluding those with long-gap and other anatomical forms that are often known to have a negative impact on outcomes, allows for a homogeneous group of patients. Finally, the systematic implementation of the most recent standards for paraclinical examinations enabled the optimisation of the presented results. Nevertheless, further prospective studies remain mandatory to validate the results presented here.

## 5. Conclusions

In this series of patients with type III EA, nutritional status and pulmonary function were impaired at the age of 7 in 25.5% and 36.7% of cases, respectively. Restrictive impairment was identified in 31% of cases, while obstruction was observed in only 9.7%. Our results suggest that particular attention should be paid to the monitoring of Caucasian patients with heart defects, tracheomalacia, and neonatal GERD, as well as those requiring respiratory treatments and showing lower BMI at school age. These patients may be at a higher risk of developing adverse respiratory conditions, and thus require closer monitoring, including early pulmonological monitoring through PFT. It should be noted that chest wall deformities and surgical technical concerns are unlikely to have a significant impact on lung function impairment.

## Data Availability

All data produced int th present study are available upon reasonable request to the authors.

## Acknowledgements

The authors would like to express their gratitude to Pr Guillaume Podevin for his invaluable assistance in the conceptualization of this study, to Dr Brittany Hemery for her expertise in the standardization of pulmonary function tests, to the CRACMO, and to Mrs Katialine Groff, Clinical Research Associate, for her dedication in the management of this study. The CRACMO registry is also acknowledged for their assistance in the management of this study, as are Mrs Joséphine Dauchet, Océane Zaghet, Isabelle Petit, Claire Dimier, Aurélie Cazals, and Aline Joulié, Clinical Research Associates, who provided invaluable assistance with data collection.

## Declaration of Generative AI and AI-assisted technologies in the writing process

During the preparation of this work, the authors used DeepLPro online software to correct and improve the quality of English writing. After using this tool, the authors reviewed and edited the content as needed and take full responsibility for the content of the publication.

## Declaration statements

The authors have no conflicts of interest to declare.

This study was financially supported by the University Hospital Centre of Angers (grant number 49RC19_0185_2), the non-profit sector associations “Institut de Recherche en Santé Respiratoire des Pays de la Loire (IRSR-PdL)” and “Filière des Maladies rares Abdomino-THOraciques (FIMATHO)”.

A selection of these results were presented as oral presentations at the 2023 IPEG/ESPES Congress, the 2024 Congress of the French Pediatric Society (SFP) and the 2024 Congress of the French Pediatric Surgery Society (SFCP).

## Data sharing statement

The datasets used during the current study are available from the corresponding author on reasonable request.

## Abbreviations and Acronyms

BMI: body mass index
CRACMO: Centre de Référence des Affections Congénitales et Malformatives de l’Oesophage
EA: esophageal atresia
FEV1: Maximal Expiratory Volume in one second FVC: Functional Vital Capacity
GERS: gastro esophageal reflux disease GLI: Global Lung Initiative
NLF: normal lung function PFT: pulmonary function tests
PNDS: Plan National de Diagnostic et de Soins
VACTERL: Vertebral defects, Anal atresia, Cardiac defects, Tracheo-Esophageal fistula, Renal anomalies and Limb abnormalities.

---

**Supplemental data 1 :**
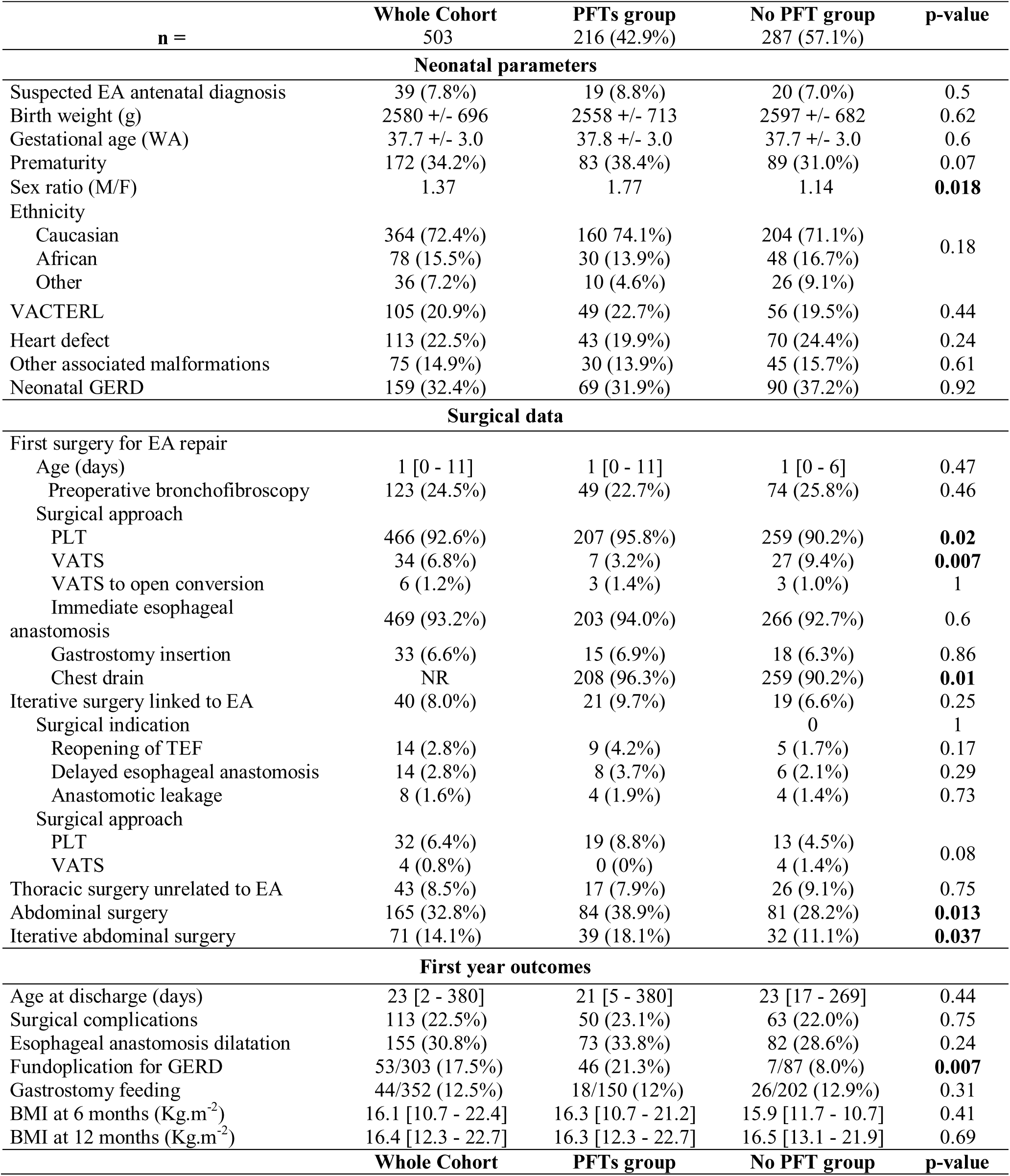
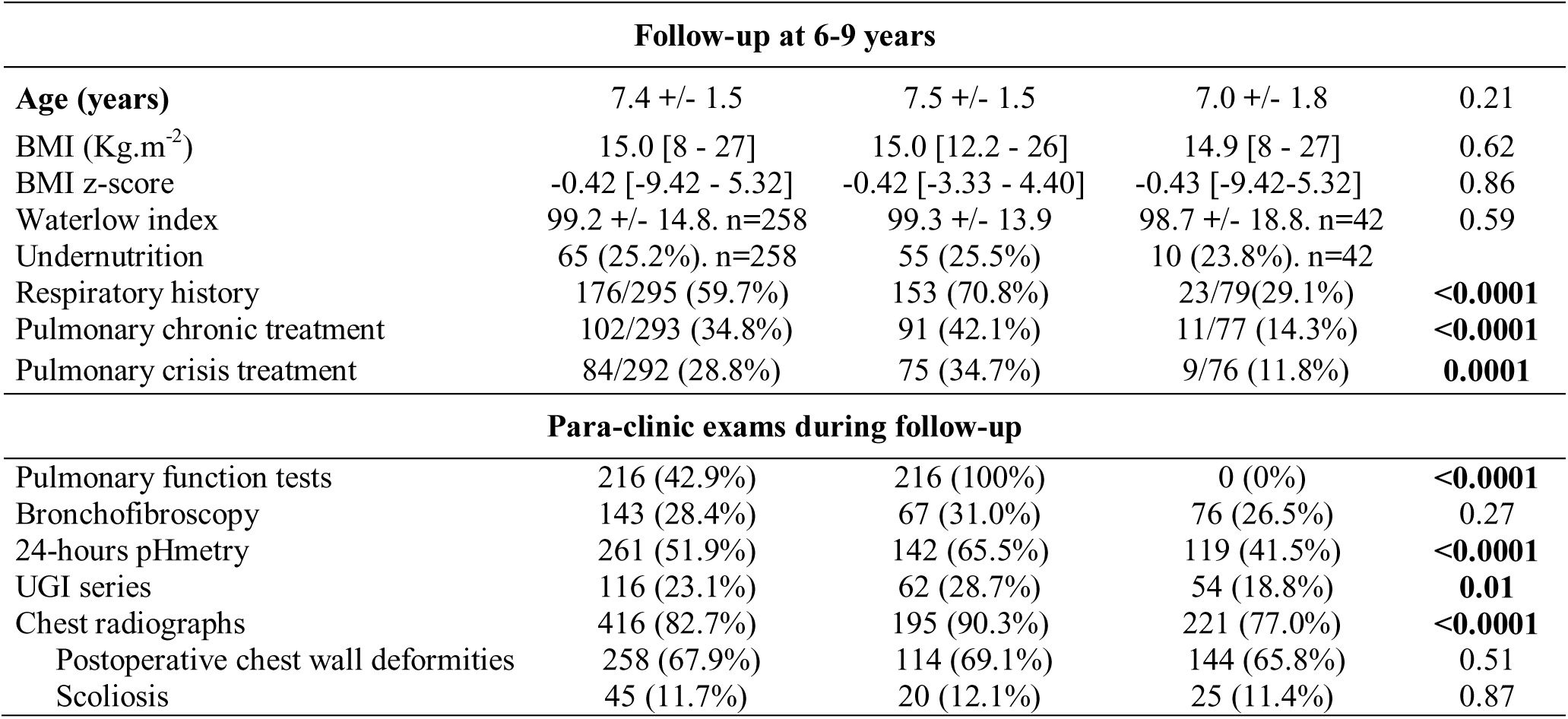
Detailed patients’ characteristics of the whole cohort of patients born with type III esophageal atresia and comparison between groups having or not pulmonary function tests at school age.

Quantitative variables are expressed as median with interquartile range or mean +/- standard deviation (SD) and qualitative data as absolute number with percentages. Statistically significant results have been expressed in bold letter. WA : weeks of amenorrhea ; GERD : gastro-esophageal reflux disease ; EA : esophageal atresia ; PLT : posterolateral thoracotomy ; VATS : video-assisted thoracoscopy; BMI : body mass index ; UGI : upper gastro-intestinal.

On the whole cohort of 503 patients, mean gestational age was 37.7 ± 3.0 WA and mean birth weight 2580 ± 696 g. The sex ratio was 1.4 in favor of boys, 172 (34.2%) patients were premature, 105 (20.9%) presented with abnormalities of the VACTERL sequence, 113 (22.5%) had congenital heart disease and 75 (14.9%) diverse other malformations. At a median age of 1 [1–2] day, surgery consisted in the closing of the tracheo-esophageal fistula in 100% of the cases, associated with esophageal reconstruction in 469 (93.2%) patients. It was preceded by a bronchofibroscopy in 123 (24.5%) cases, and was performed by a posterolateral right thoracotomy in 466 (92.6%) patients and by thoracoscopy for 34 (6.8%) patients. Forty (8.0%) patients required a second thoracic intervention for esophageal atresia, 43 (8.5%) others for cardiac or pulmonary causes and 165 (32.8%) had at least one abdominal surgery during their first year. They were discharged home after 23 [15–47] days, and 80.7% of them had proton pump inhibitors as GERD treatment.

During their follow-up and at a mean age of 7.4 ± 1.5 years, 28.4% of the patients had at least one bronchofibroscopy, 23.1% upper gastrointestinal series, 51.9% a 24h-pHmetry and 76.3% chest radiographs. Mean BMI was 15.6 ± 2.5 and Waterlow index 99.2 ± 14.8%, with 17.9% of the patients presenting signs of GERD, 22.3% currently treated by PPI, and 17.5% having had a fundoplication. Chest wall deformities were found in 273 out of 384 (71.1%) radiographs, including 258 (67.9%) intercostal space abnormalities resulting from surgery, 45 (11.7%) scoliosis and 23 (6.0%) vertebral malformations. A respiratory history was found in 60% of the patients, which required chronic treatments in 34.8% (102/293 available data) and had had some crisis treatment in the past six months in 28.8% of the cases

Comparison of the groups of patients with (n = 216) and without (n = 287) interpretable PFT found no significant difference in terms of perinatal data and anthropometric data at school age. There were some minor differences in surgical management between the two groups. Patients without PFT had more thoracoscopies (9.4% vs 3.2%, p = 0.007) and fewer pleural drains (90.2% vs 96.3%, p = 0.01) than those with PFT. There was a notable reduction in the number of abdominal surgeries for this group (28.2% vs 38.9%, p = 0.01). However, there was no significant difference in the frequency of iterative thoracic interventions or surgeries related to EA. At follow-up, patients with PFT also underwent more paraclinical examinations for respiratory or digestive purposes, although the results were similar.

## Notes

### Competing Interest Statement

The authors have declared no competing interest.

### Funding Statement

This study was funded by the University Hospital Centre of Angers (grant number 49RC19_0185_2), the non-profit sector association "Institut de Recherche en Sante Respiratoire des Pays de la Loire (IRSR-PdL)" and the "Filiere des Maladies rares Abdomino-THOraciques (FIMATHO)".

### Author Declarations

Ethics committee of the University Hospital Centre of Angers (France) gave ethical approval for this work.

